# Does sampling saliva increase detection of SARS-CoV-2 by RT-PCR? Comparing saliva with oro-nasopharyngeal swabs

**DOI:** 10.1101/2020.07.26.20158618

**Authors:** Ozlem Akgun Dogan, Betsi Kose, Nihat Bugra Agaoglu, Jale Yildiz, Gizem Alkurt, Yasemin Kendir Demirkol, Arzu Irvem, Gizem Dinler Doganay, Levent Doğanay

## Abstract

The gold standard method in the diagnosis of SARS-CoV-2 infection is the detection of viral RNA in nasopharyngeal sample by RT-PCR. Recently, saliva samples has been suggested as an alternative due to being fast, reliable and non-invasive, rather than nasopharyngeal samples. We compared RT-PCR results in nasopharyngeal, oro-nasopharyngeal and saliva samples of COVID-19 patients. 98 of 200 patients were positive in RT-PCR analysis performed before the hospitalization. In day 0, at least one sample was positive in 67% of 98 patients. Positivity rate was 83% for both oro-nasopharyngeal and nasopharyngeal samples, while it was 63% for saliva samples (*p*<0.001). On day 5, RT-PCR was performed in 59 patients, 34% had at least one positive result. The positivity rate was 55% for saliva and nasopharyngeal samples, while it was 60% for oro-nasopharyngeal samples. Our study shows that the sampling saliva does not increase the sensitivity of RT-PCR tests at early stages of infection. However, on 5th day, viral RNA detection rates in saliva were similar to nasopharyngeal and oro-nasopharyngeal samples. In conclusion, we suggest that, in patients receiving treatment, virus presence in saliva, in addition to the standard samples, is important to determine the isolation period and to control the transmission.

## Introduction

The SARS CoV-2 infection, which has influenced the world since the end of 2019, causes serious problems in both health and socio-economic fields.^1,2^ Currently, it has affected approximately 6 million people over more than 200 countries.^3^ SARS-CoV-2 infection can cause serious lower respiratory tract infections that can be fatal in some patients, however, many individuals remain asymptomatic during the infection.^4,5^ Especially asymptomatic individuals have been a major factor in increasing the transmission rate of the disease and evolving it into a pandemic.

COVID-19 diagnosis is based on clinical findings, besides, the detection of the virus in patients’ specimens is of great importance in terms of monitoring the disease, guidance of treatment, and infection control,^6^ Quarantine, which is launched all over the world urging people to stay home, is considered as the only way to reduce transmission, but it started to cause serious social and economic problems due to loss of labor.^7,8^ This situation reveals the need for a fast, reliable, easily applicable, and non-invasive test that quickly identifies infected individuals to be isolated. Currently, gold standard method in the diagnosis of SARS-CoV-2 infection is the detection of viral RNA in the nasopharyngeal swab sample by Real Time Polymerase Chain Reaction (RT-PCR) analysis.^9^ The most important disadvantage of this method is the presence of limited trained personnel available in sampling during the outbreak. High risk of nosocomial infections that such personnel are exposed to is an additional obstacle. Moreover, in terms of patients, discomfort experienced, especially, in repeated tests is the most frequently reported problem.

The abovementioned disadvantages have led the researchers to study on non-invasive, easy to self-applicable sampling methods for massive screening.^10–14^ As the main source of transmission of SARS-CoV-2 infection is salivary droplets, viral RNA RT-PCR in saliva samples have been suggested as possible alternative testing for diagnosis.^15^ The main advantages of saliva sampling are self-availability, no need for specialized staff and the comfort of the procedure. However, although turnaround time and self-applicability of the tests are important, the reliability of the sampling method to be chosen should also be tested very well. It is obvious that the use of tests with a high false negativity rate will adversely affect the course of the pandemic.

Here, we compared the results of RT-PCR in nasopharyngeal, oro-nasopharyngeal, and saliva samples in patients diagnosed with COVID-19 to investigate their possible relationships with clinical findings.

## Methods

### Patients

A cross-sectional study was conducted in repurposed Genomic Laboratory (GLAB), Umraniye Teaching and Research Hospital, in Istanbul, with a total of 200 consecutive patients who met the possible case definition for COVID-19 and hospitalized with moderate-severe disease.^16^ According to the diagnostic algorithm provided by The Turkish Ministry of Health, the possible cases were defined as those who presented with;

*History of fever or acute respiratory symptoms,

and

*Travel history from an endemic area of COVID-19 within 14 days,

or

*History of contact with an individual who was confirmed or suspected having COVID-19,

or

*Presence of hospitalization requirement due to respiratory tract infection.

Within the scope of the present study, after hospitalization saliva, oro-nasopharyngeal and nasopharyngeal samples were taken from all 200 patients within the first 24 hours, and it was defined as day 0 sample. On day 5, patients were resampled.

Demographic characteristics, symptoms at presentation, comorbid diseases, and clinical findings during hospitalization were collected for each patient. All subjects provided informed consent, and the study was approved by the ethics committee of Umraniye Teaching and Research Hospital (B.10.1.THK.4.34.H.GP.0.01/167)Patients admitted to ICU, not giving consent to study, incapable of providing saliva sample and patients under age of 18 were excluded.

### Sample Collection

In all patients, an oro-nasopharyngeal sample was taken with a cotton swab used for the viral RT-PCR test before hospitalization as a standard diagnostic approach. In the scope of our study, oro-nasopharyngeal samples were taken with cotton swab and nasopharyngeal samples with dacron swab. Details of the sampling processes are given in Figure 1. Before the saliva collection, participants were given brief explanations about the difference between saliva and sputum, then they were asked to give saliva samples prior to other samples by a drooling technique. They spit approximately 1 mL into the falcon tubes containing the viral transport medium (VTM, Innomed VTM001) used in standard sampling. Single trained healthcare professional took samples to avoid possible variations in the collection technique. All samples were transferred to our laboratory within 1 hour of sampling stored in the refrigerator, and RT-PCR was performed on the day they were collected.

**Figure 1:**
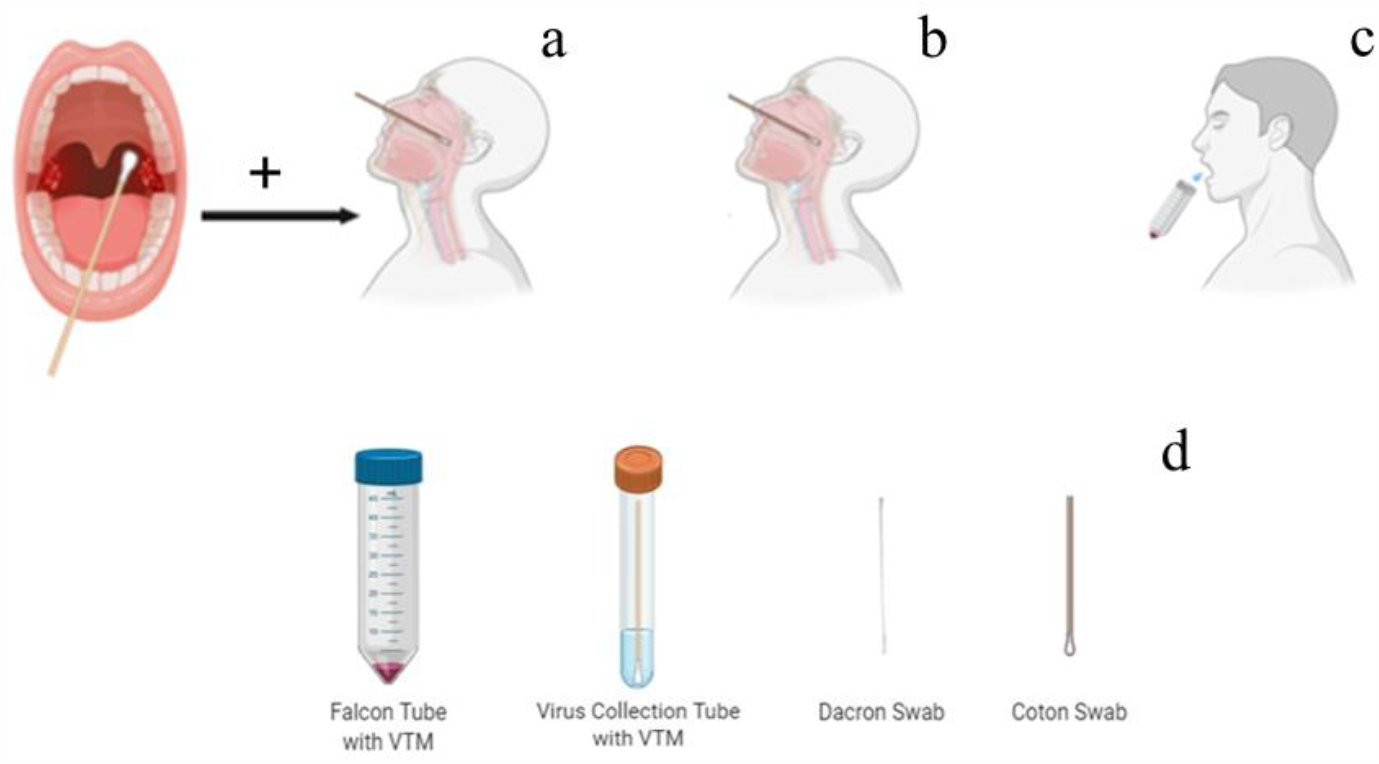
Schematic representation of the sampling methods used in the study (1a: Oronasopharyngeal sampling, 1b: Nasopharyngeal sampling, 1c: Saliva sampling, 1d:Tube and swab types) (This figure was designed by using BioRender program that is licensed)

### RT-PCR Workflow

*ORF1ab* and *N gene* of SARS-CoV-2 were targeted for the diagnosis of the infected patients. For this purpose, The Direct Detection of SARS-CoV-2 Detection Kit was used (Coyote Bioscience Co., Ltd). The kit procedure was based on the detection of the conserved region of ORF1ab coated with a pair of specific primers and a fluorescently labelled probe, and the N gene of SARS-CoV-2 by the RT-PCR method. Since this kit did not need a separate RNA extraction, the samples in the VTM medium had briefly vortexed and taken directly into the study. Biorad CFX 96 Real-Time PCR systems were used. FAM channel for *ORFlab* gene, ROX channel for *N gene*, and HEX channel for internal *RNase P* gene of human control were selected. These channels should have the logarithmic growth period with the Ct value ≤29, for a positive result.

### Interpretation of Results

In the RT-PCR results of the samples, the internal RNase P gene was positive for all samples. If both the ORFlab and the N gene were positive, the result was considered as presumptive positive. If both the ORFlab and the N gene were negative, the result was considered as presumptive negative. If one of them was positive and the other was negative, the test for this sample was repeated. If the same result was achieved again, a new nasopharyngeal and oropharyngeal swab was requested.

### Statistical Analysis

SPSS.22 package program (IBM Company, Armonk, NY, USA) was used for statistical analysis. Mean, median and standard deviation were used for descriptive statistical information. Categorical variables were analysed with chi square test. The results of the tests performed in three different samples on the 0th and 5th days were compared among themselves using the Cohran Q test. All calculated P values are double sided and for significant statistical results, *p* <0.05 was accepted.

## Results

98 of 200 (49%) patients in the study group were positive in RT-PCR analysis performed on the samples taken as a standard diagnostic procedure before the hospitalization. The clinical and demographic characteristics of the patients with positive results are presented in Table 1. Although 102 patients met the possible case definition, the viral RT-PCR tests on admission were negative (Figure 2).

**Table 1:** The clinical and demographic characteristics of the patients.

|  | Patients with positive RT-PCR results<br>n=98 | Patients with negative RT-PCR results<br>n=102 | Total<br>n=200 | p value |
| --- | --- | --- | --- | --- |
| **Age (yrs, mean ±SD)** | 55.8(±15.8) | 54.1(±16.4) | 54.9(±16.1) | 0.434 |
| **Sex M/F** | 48/50<br>49% | 58/44<br>57% | 106/94<br>53% | 0.264 |
| **Symptoms of presentation** |  |  |  |  |
| -Cough | 68<br>69% | 52<br>51% | 120<br>60% | 0.008 |
| -Myalgia | 72<br>73% | 38<br>37% | 110<br>55% | 0.000 |
| -Fever | 44<br>45% | 36<br>35% | 80<br>40% | 0.166 |
| -Dyspnea | 42<br>43% | 47<br>46% | 89<br>45% | 0.647 |
| -Loss of smell&taste | 9<br>9% | 4<br>4% | 13<br>7% | 0.131 |
| **Thorax CT (%)** |  |  |  | 0.001 |
| -Normal | 8<br>8% | 22<br>22% | 30<br>15% |  |
| -Mild Infiltration | 53<br>54% | 59<br>58% | 112<br>56% |  |
| -Moderate Infiltration | 32<br>33% | 17<br>17% | 49<br>25% |  |
| -Severe Infiltration | 5<br>5% | 4<br>4% | 9<br>5% |  |
| **Thorax CT Moderate & Severe vs Normal & Mild** | 37<br>38% | 18<br>18% | 55<br>28% | 0.001 |

**Figure 2:**
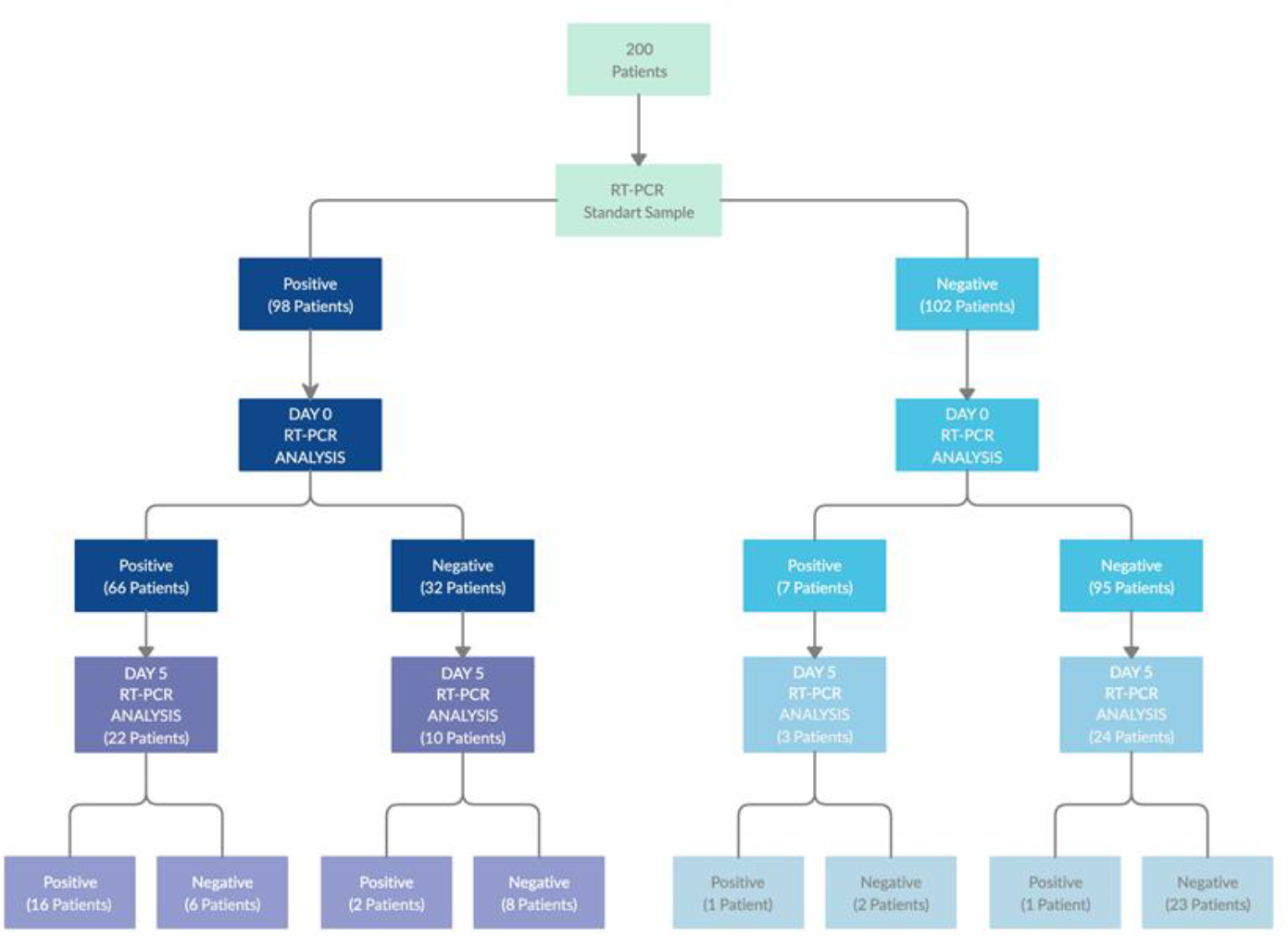
Study workflow diagram.

At day 0 RT-PCR analysis, at least one sampling method showed positivity in 66 of the above mentioned 98 patients. The sensitivity rate was observed as 55/66 (83%) for both oro-nasopharyngeal and nasopharyngeal samples, while it was 35/66 (63%) for saliva samples (Table 2) and the difference of the sensitivity rates among the sampling methods was statistically significant (*p*< 0.001) (Figure 3a) The mean C*t* values determined for FAM and ROX were shown in Figure 4.

**Table 2:**
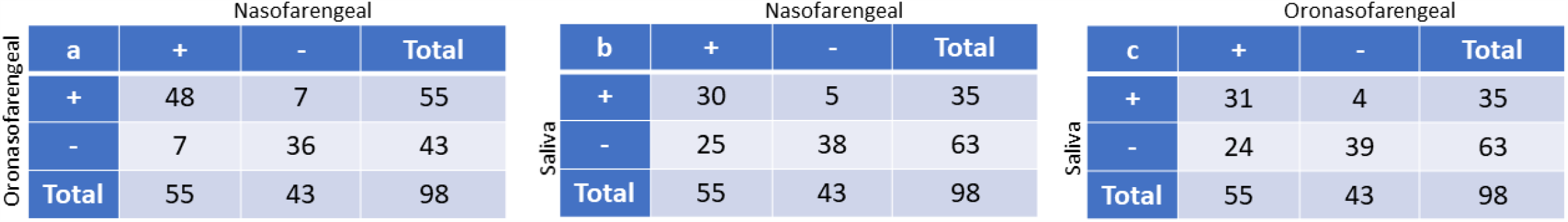
RT-PCR study results of saliva, oronasopharyngeal and nasopharyngeal samples on day 0 in 98 patients who were found positive in RT-PCR analysis before hospitalization.

**Table 3:**
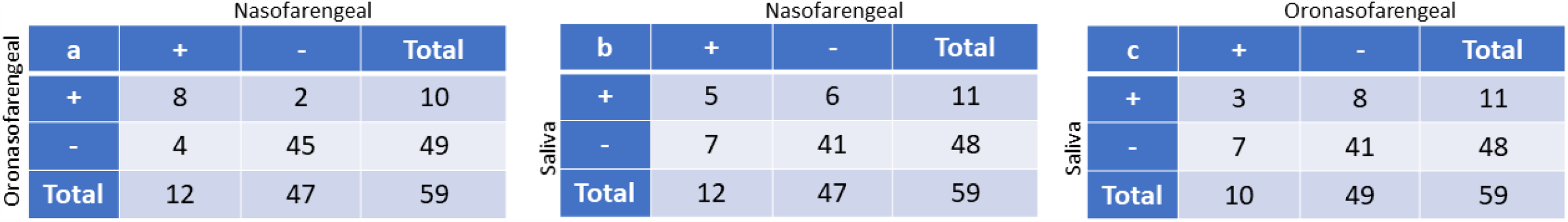
RT-PCR study results of saliva, oro-nasopharyngeal and nasopharyngeal samples on day 5 in 59 patients. We would like to remind that of 59 patients, 20 (34%) had at least one positive result in saliva or swab sample.

**Figure 3:**
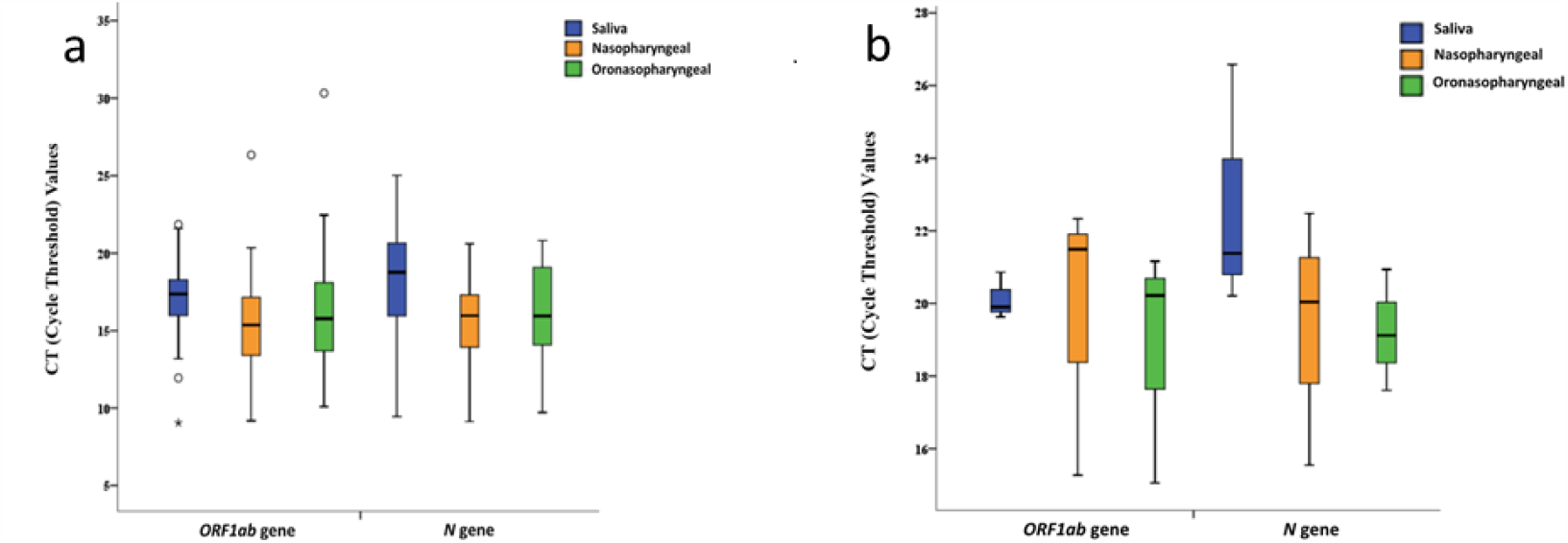
Sensitivity rate of each sampling methods in day 0 and 5 RT-PCR analysis.

**Figure 4:**
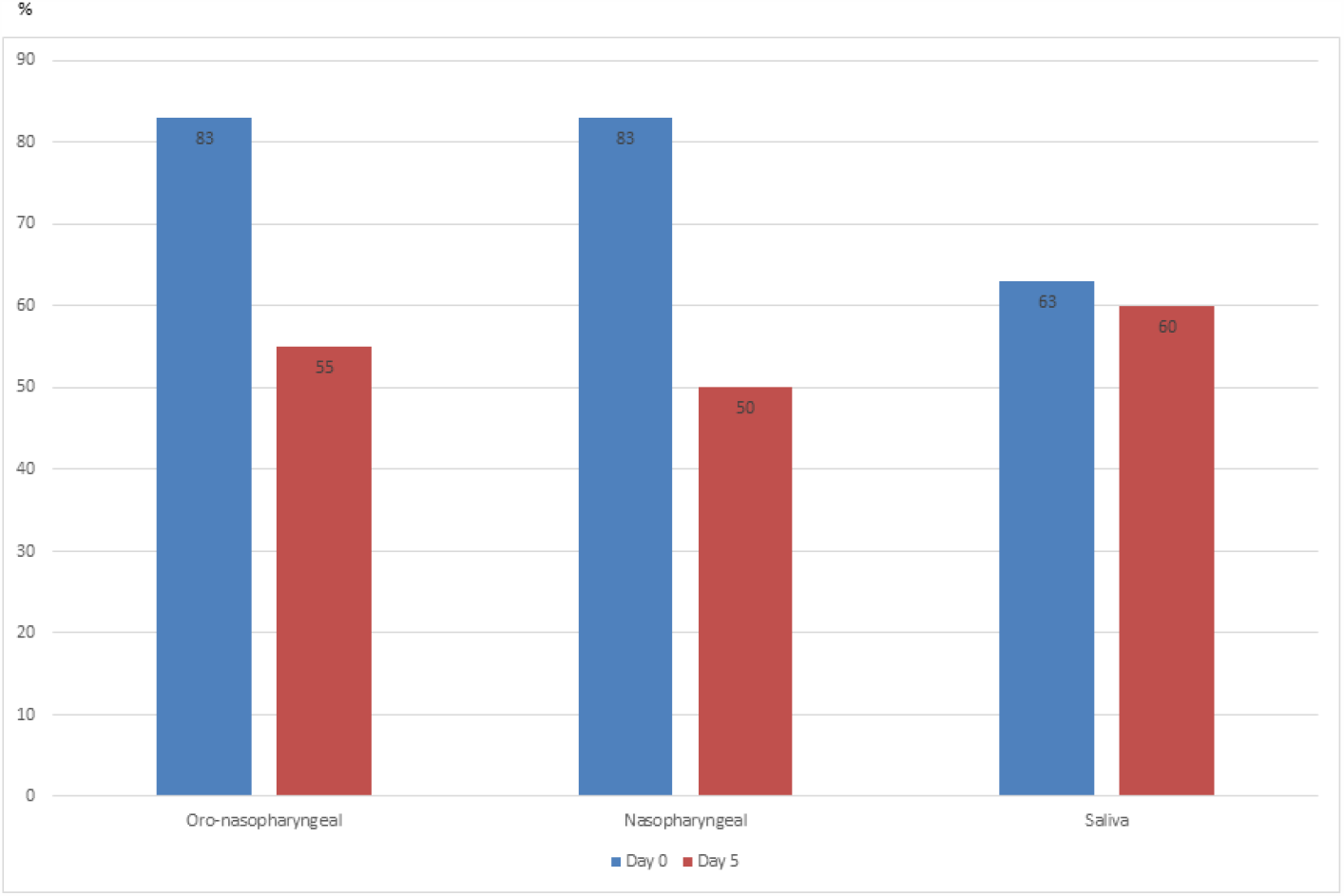
Graphical representation of FAM and ROX Ct values detected in saliva, nasopharyngeal and oronasopharyngeal samples in day 0 (a) and 5 (b) RT-PCR studies.

On day 0, 6 (9%) patients had positive result only in nasopharyngeal swab sample, 5 (8%) was only positive in oro-nasopharyngeal sample and additional 3 (5%) patients had a positive test results only in saliva samples. Those three patients who had only saliva positivity had a history of COVID-19 positive household contact and had mild involvement in thorax CT.

In the RT-PCR analysis on day 0, of 102 patients with a previous negative result, 95 remained as negative, while in 7 (6·9%) patients positive RT-PCR results obtained in at least one sample type (Figure 2). While the oro-nasopharyngeal sample was positive in all 7 patients, nasopharyngeal samples were positive in 3. There was only one patient whose saliva sample was positive. Oro-nasopharyngeal and nasopharyngeal samples in this patient were also positive.

On day 5, we were able to collect samples in 59 of 200 (30%) patients. Of 59 patients, 20 (34%) had at least one positive result in saliva or in one of the swab samples. The sensitivity rate was determined as 11/20 (55%) for both saliva and nasopharyngeal samples, while it was 12/20 (60%) for oro-nasopharyngeal samples (Figure 3b). In the statistical analysis, there was no difference between the sample types in terms of the sensitivity (*p*=0.368). The mean C*t* values determined for FAM and ROX were shown in Figure 4. On day 5, there were 5 patients who showed RT-PCR positivity only on the saliva sample. These patients had longer prior history with positive RT-PCR results in oro-nasopharyngeal samples, home-treated with hydroxychloroquine for five days, clinically deteriorated with radiological findings, tested positive again in oro-nasopharyngeal samples and hospitalized.

## Discussion

Our study demonstrates that saliva sampling did not improve diagnostic sensitivity in patients who had a negative result in initial testing before hospitalization. Our results also revealed that at the early stages of the infection, saliva sampling had a lower sensitivity to detect viral RNA compared to other sampling methods. In contrast to our findings, three recent studies suggest that the results of RT-PCR analysis in saliva samples are compatible with the results of the nasopharyngeal and oro-nasopharyngeal samples^11,13,14^. These studies had either limited number of patients (23 and 12 patients) or limited number of RT-PCR positive patients (21 RT-PCR positive patients). In this study we consecutively recruited 200 inpatients who presented with clinical signs compatible with COVID-19 and at initial testing 98 of them revealed SARS-COV-2 RT-PCR positivity. In this study the sensitivity rate for saliva samples was 63% and this was significantly lower than nasopharyngeal and oro-nasopharyngeal swabs. However, our data on the 5th day showed that the viral RNA detection rates in saliva samples were similar to those of nasopharyngeal and oro-nasopharyngeal samples. Two recent studies supported our results and showed that SARS-CoV-2 detection from saliva was more consistent during extended hospitalization and recovery^11,14^. Thus, we suggest that RT-PCR analysis in saliva samples may be beneficial when afollow up test beyond day 5 is needed especially in hospitalized patients (e.g. before discharge). Compared to nasopharyngeal swabs, sampling saliva causes less discomfort in patients and reduces risk for nosocomial infection among healthcare workers as patients can give saliva samples by themselves.

On day five, out of 59 patients, 5 (8%) had detectable viral RNA only in saliva sample. The fact that all of these patients were using hydroxychloroquine prior to hospitalization, this may have hindered the detection of the virus in nasopharyngeal and oro-nasopharyngeal samples. With this result, we suggest that taking saliva samples along with the standard method before ending the isolation in individuals treated with hydroxychloroquine can be effective in minimizing contamination in the community by reducing the false negativity rate. In addition, we recommend saliva sampling in patients who show progression despite at-home treatment. Such an approach would increase the detection of positive cases, thereby enabling more accurate planning of treatment, follow-up, and subsequent discharge.

One of the most important issues affecting false negative rates in standard nasopharyngeal sampling is the use of inappropriate techniques. In our study, taking all samples by single trained healthcare staff is the strength of our study and enabled us to achieve a safer result. The most important limitation of our study is that no study has been performed in asymptomatic individuals and mild cases that do not require hospitalization. Therefore, it was not possible to make a comment about whether saliva samples can be used in screening.

In conclusion, our study shows that the saliva sample is not as sensitive as standard nasopharyngeal swabs in determining viral RNA and it does not improve detection rate in PCR negative patient group. However, in the later stage of the disease, RT-PCR test from saliva samples might help detecting deteriorating patients or determining the isolation period more effectively after treatment.

## Data Availability

All data generated or analysed during this study are included in this article.

## Acknowledgements

We thank to Murat Kaya for technical assistance

